# Prevalence and pattern of adverse events following COVID 19 vaccination among adult population in Sokoto metropolis, northwest, Nigeria

**DOI:** 10.1101/2022.11.01.22281793

**Authors:** Habibullah Adamu, Sufyanu Lawal, Ishaka Alhaji Bawa, Akilu Muhammad Sani, Adamu Ahmed Adamu

## Abstract

**Background:** COVID-19 still poses a major public health challenge worldwide and vaccination remains one of the major interventions to control the disease. Different types of vaccines approved by the World Health Organization (WHO) are currently in use across the world to protect against the disease. As all vaccines are associated with some adverse reactions (ARs), this study assessed the prevalence and pattern of adverse events following immunization (AEFI) after receiving COVID-19 vaccine among the adult population in Sokoto metropolis, North-west, Nigeria

**Methods:** We conducted a cross-sectional study among 230 adults in Sokoto metropolis who received COVID-19 vaccine. Data was collected using a structured questionnaire administered via personal phone calls to respondents who were selected via a systematic sampling technique. For data analysis, IBM SPSS version 25.0 was used.

**Results:** The Majority of the participants [183 (79.7%)] experienced AEFI. The most common adverse events were body weakness [157(85%)], fever [111(60.3%)] and headache [103(56%)]. Up to half of the respondents that experienced AEFI said it occurred within minutes and a few hours, whereas 75 (40.8%) said it was within 2-3 days. Up to 66.3 of the adverse reactions were mild and lasted between a few hours (37.5%) and one day (31.5%); however, 15.2% of the respondents had severe reactions of which 22.7% were admitted to a health facility. The development of AEFI was linked to the absence of an underlying medical condition, a previous history of AEFI, and a history of drug reaction.

**Conclusion:** The majority of respondents reported adverse events following COVID-19 vaccination, with body weakness, fever, and headache being the most common AEFIs. The underlying medical condition as well as a history of adverse drug reactions were predictors of the development of adverse reactions following COVID-19 vaccination. Service providers at each COVID-19 vaccination point should always take the time to explain to vaccine recipients that adverse reactions are possible; however, they should reassure them that most ARs resolve within a few hours to a few days.

## Introduction

Severe acute respiratory syndrome the coronavirus 2 (SARS-CoV-2) is the causative virus for the coronavirus disease 2019 (COVID-19) ongoing pandemic. SARS-CoV-2 first emerged in late 2019 in Wuhan (Hubei, China) and hastily became a global threat, affecting over 200 countries.^1,2^ The pandemic has resulted in a devastating impact worldwide, which prompted the need for mitigation policies to contain it. Till date, there is no known cure for COVID-19, thus control measures are largely preventive in nature, and these include pharmacological and non-pharmaceutical interventions (NPIs) such as regular use of face masks, use of hand sanitizers, social distancing, travel restrictions, partial or complete lockdowns, and use of vaccines.^2^

Because of the high disease burden of SARS-CoV-2, the development and manufacture of COVID-19 vaccines has been occurring at an unprecedented pace. Different vaccines against COVID-19 have now been approved for use by the World Health Organization (WHO); these vaccines include Oxford-Astrazeneca, Moderna, Pfizer-BioNtech, Johnson & Johnson and many others.^3^ Different technologies have been adopted in the development of these vaccines and many are emerging with different mechanisms of action.^3^ Findings from clinical trials have shown the vaccines to be effective and safe for use among humans;^4^ The efficacy of Pfizer and Moderna vaccines in preventing disease or severe disease is 95–87.5% and 94.5–100%, respectively, whereas the efficacy of Astra-Zeneca and Janssen is about 70% and 65%, respectively.^5^ Since the initiation of the COVID-19 vaccination programs in many countries, concerns have grown about adverse events following immunization (AEFI), especially with respect to COVID-19 vaccines.^6,7^

To achieve the goal of global eradication of COVID-19, at least 70% of all persons must receive the COVID-19 vaccine.^8^ As at 3^rd^ October 2022, up to 68% of global population has received at least one dose of COVID-19; in Latin America, Cuba had the highest vaccination coverage rate (95.1%), in Europe, Portugal had the highest rate (94.8%).^9^ Nigeria aimed to vaccinate 40% of its over 200 million people by the end of 2021 and hopes to reach the 70% vaccination threshold for eliminating COVID-19 by the end of 2022. As of October 15, 2022, a total of 44,687,220 eligible adults (i.e. 40% of the 111,765,503 target population) had been fully vaccinated against COVID-19 nationwide; 57,433,884 people (51.4%) had only received the first dose.

Osun state has the highest proportion of those fully immunized (89%) whereas Bayelsa and Rivers states have the lowest proportion (5.3%); in Sokoto state, 44.3% of the target population have been fully vaccinated.^11^

An adverse event following immunization is any untoward medical occurrence which follows immunization and does not necessarily have a causal relationship with the use of the vaccine; the adverse event may be any unfavourable or unintended sign, abnormal laboratory finding, symptom or disease.^12^ Because severe reactions following immunization are rare, several countries pool their AEFI data in a common global database; the database is managed by the WHO Programme for International Drug Monitoring. It has been observed that a significant number of severe AEFI are not true vaccine reactions but rather, coincidental occurrences of health events or the anxiety associated with the receipt of a vaccine.^13^

Like with most vaccines, most local or systemic AEFI to COVID-19 vaccines are mild to moderate in severity, and the overall frequency of the AEFI varies with the age of the recipient and type of vaccine given, among others. Reports suggest that the rate of adverse reactions (ARs) to the Oxford-Astrazeneca vaccine is higher than that of the Pfizer-BioNtech vaccine. However, the ARs to the Oxford-Astrazeneca are less frequent in the older age group.^14,15^

Several adverse events have been reported globally following the reception of different vaccines.^16^ In a systematic review of studies conducted on the efficacy of COVID-19 vaccines across the globe, it was found that the proportion of individuals who had an adverse reaction within 28 days after vaccination was lower than 30%. Most of the adverse reactions were mild to moderate and resolved within 24 hours after vaccination; pain and tenderness at the injection site were the most common local adverse reactions, whereas body weakness, pain and fever were the most frequent systemic adverse reaction.^4^ In India, a study involving 1826 recipients of COVID-19 vaccines showed that 29.8% of them reported at least one of the AEFI; no severe adverse event was reported, and only 1.6% of them had moderate AEFI. Pain at the injection site (14.6%), fever (9.7%), and myalgia (5.9%) were the most common adverse events reported by the recipients. The AEFI incidence was higher in the first dose (38.1%) when compared to the second dose (26.4%).^17^ In Poland, a study among healthcare workers showed that 78.9% had at least one local reaction and 60.7% had at least one systemic reaction; pain at the injection site was the most prevalent local reaction (76.9%), while fatigue (46.2%), headache (37.7%) and muscle pain (31.6%) were the most common systemic reactions.^18^

In Africa, an online survey conducted among participants from 22 African countries revealed that 80.8% of the respondents reported an adverse event following COVID-19 vaccination.^16^ In Togo, at least one adverse event after receiving COVID-19 vaccine was reported among 71.6% of participants in a study; the most commonly reported adverse events were pain on injection site (91.0%), asthenia (74.3%), headache (68.7%), soreness (55.0%), and fever (47.5%). The prevalence of severe adverse events (SAEs) was 23.8%, with being a female under the age of 50 being more associated with the occurrence of SAEs ^19^.

In Nigeria, the pattern of adverse reactions to COVID-19 vaccines is similar to what has been observed in other countries; a cross-sectional study conducted in Rivers state showed that 50.5% of recipients of COVID-19 vaccine reported at least one adverse event post-vaccination, out of which 4.6% were said to be severe. The most common ARs were fever (73.0%), pain at the injection site (41.2%), fatigue (33.3%), body aches (17.5%) and headache (13.8%).^20^ In Sokoto state, up to 1,252,502 people have been fully vaccinated against COVID-19 ^11^. However, there is no available data on AEFI experienced by recipients in the state. As fear of side effect has been mentioned as one of the reasons for COVID-19 vaccine hesitancy,^21^ it is therefore, important to determine the prevalence and pattern of adverse reactions experienced by recipients of the vaccines in Sokoto state; this forms the basis for this study.

## Materials and methods

### Study Area

This study was carried out at Usmanu Danfodiyo University Teaching Hospital (UDUTH), Primary Healthcare Center (PHC) Arkilla and Specialist Hospital, all within the Sokoto metropolis.

UDUTH is a tertiary hospital that provides both health and academic services to Sokoto, Kebbi and Zamfara States and other parts of Nigeria, including some parts of neighbouring Niger and Benin Republics. It offers clinical services in the form of preventive, diagnostic, curative and rehabilitative services. The Centre for Advanced Medical Research and Training (CAMRET), which has been approved by the Nigeria Centre for Disease Control (NCDC) to carryout COVID-19 test is located within the hospital premises; UDUTH currently provides COVID-19 vaccination services using Pfizer BioNtech, Moderna, Oxford Astrazenica, and Johnson and Johnson vaccines. Specialist Hospital Sokoto and PHC Arkilla also provide COVID-19 vaccination services and they are all located within Sokoto metropolis.

The study was conducted among the adult population who received COVID-19 vaccine within Sokoto metropolis. Only individuals who had received at least one dose of COVID-19 vaccine were included in the study. All recipients of COVID-19 vaccine whose phone numbers were not retrievable in the COVID-19 register at the respective hospitals were excluded

A cross-sectional study design was used and the sample size was estimated using the Cochrane formula for estimating sample size in descriptive study.^22^

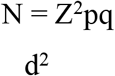 Because the total population of those who received COVID-19 vaccine at the time of the study was less than 10,000, the formula for estimating sample size for a finite population (n_f_) was used to estimate the optimal sample size.^22^

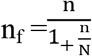

n_f_ = minimum sample size for a finite population

n= minimum sample size for infinite population

N= Total population of adults who received COVID-19 from the 3 selected health facilities = 1300

Using the above two formulae, sample size of 207 participants was estimated. A response rate of 90% was anticipated, thus the final sample size (n_f_) was adjusted to 230, by dividing the calculated sample size (207) by the anticipated response rate (0.9); therefore, 230 study participants were enrolled into the study UDUTH, Specialist Hospital Sokoto and PHC Arkilla were selected out of the health facilities that provide COVID-19 vaccination services in Sokoto metropolis using simple random sampling technique. From each of the three selected HFs, systematic sampling technique was used to select participants; the COVID-19 register in each of the vaccination points was used as sampling frame. Sampling interval for selection of participants in each HF was calculated by dividing the total number of COVID-19 vaccine recipients in the register by the sample size for that HF.

A structured, pretested questionnaire was used to obtain relevant information from the study participants. The questionnaire had four sections as follows:

Section A: Sociodemographic profile of respondents

Section B: Prevalence and pattern of AEFI following COVID-19 vaccination

Section C: Factors associated with adverse reactions following COVID-19 vaccination

The questionnaire was pretested among adult men and women who received COVID-19 vaccines from PHC Yar Akija. Necessary amendments were made thereafter, and the instrument was found valid.

Personnel used for data collection comprised the researchers with the help of three research assistants.

Data collection was done using the questionnaire described above. The questionnaire was designed and deployed to a web-based account created on https://kobo.humanitarianresponse.info. The deployed questionnaire was then accessed by downloading Open Data Kit (ODK) app on Android devices and was used for data collection. Data was collected by administering the questionnaire via personal phone calls to all the selected participants whose phone numbers could be retrieved from the COVID-19 register. Before administering each questionnaire, the research personnel explained the purpose of the research to the respondent and sought their consent to participate.

Data was retrieved from the ODK server, exported to Microsoft Excel 2016 and then transferred to IBM SPSS version 23 software for analysis. Categorical variables were analyzed and presented as frequencies and percentages, while quantitative variables were analyzed and presented as summary measures in form of measures of central tendency and their corresponding measures of dispersion. The Pearson chi-square test was used to determine the factors associated with adverse reactions following COVID-19 vaccination. A binary logistic regression model was used to identify factors that predicted AEFI following COVID-19 vaccination. The Level of statistical significance for all inferential statistical analysis was set at 5% (p<0.05)

Approval to conduct the study was obtained from the Research Ethics Committee of the Ministry of Health, Sokoto State, Nigeria and Usmanu Danfodiyo University Teaching Hospital, Sokoto. Prior to recruitment, verbal informed consent of all study participants was sought.

## RESULTS

A total of 230 questionnaires were administered, and all 230 were completed, retrieved, and analyzed, yielding a response rate of 100%.

**Table 1:** Socio demographic profile of the respondents.

| **Variable** | **Frequency(n=230)** | **Percentage (%)** |
| --- | --- | --- |
| **Age group ( years)** |  |  |
| ≤20 | 2 | 0.9 |
| 20-39 | 129 | 55.8 |
| 40-59 | 78 | 33.8 |
| 60 and above | 22 | 9.5 |
| **Sex** |  |  |
| Male | 138 | 60.2 |
| Female | 92 | 39.8 |
| **Religion** |  |  |
| Christianity | 10 | 4.3 |
| Islam | 220 | 95.7 |
| **Tribe** |  |  |
| Hausa/Fulani | 215 | 93.1 |
| Igbo | 8 | 3.9 |
| Yoruba | 7 | 3.0 |
| **Marital status** |  |  |
| Married | 175 | 75.8 |
| Divorced | 7 | 3.0 |
| Widow | 5 | 2.2 |
| Single | 43 | 19.0 |
| **Occupation** |  |  |
| Health worker | 31 | 13.4 |
| Businessman/businesswoman | 101 | 43.7 |
| Self employed | 51 | 22.5 |
| Farmer | 8 | 3.5 |
| Student | 39 | 16.9 |

Majority of respondents were in the age group of 20-39 years of age (55.8%) with others between 40-59 years of age (33.8%). Most of the respondents were males (60.2%), married (75.8%) and practice Islam as religion (95.7%).

**Figure 1.**
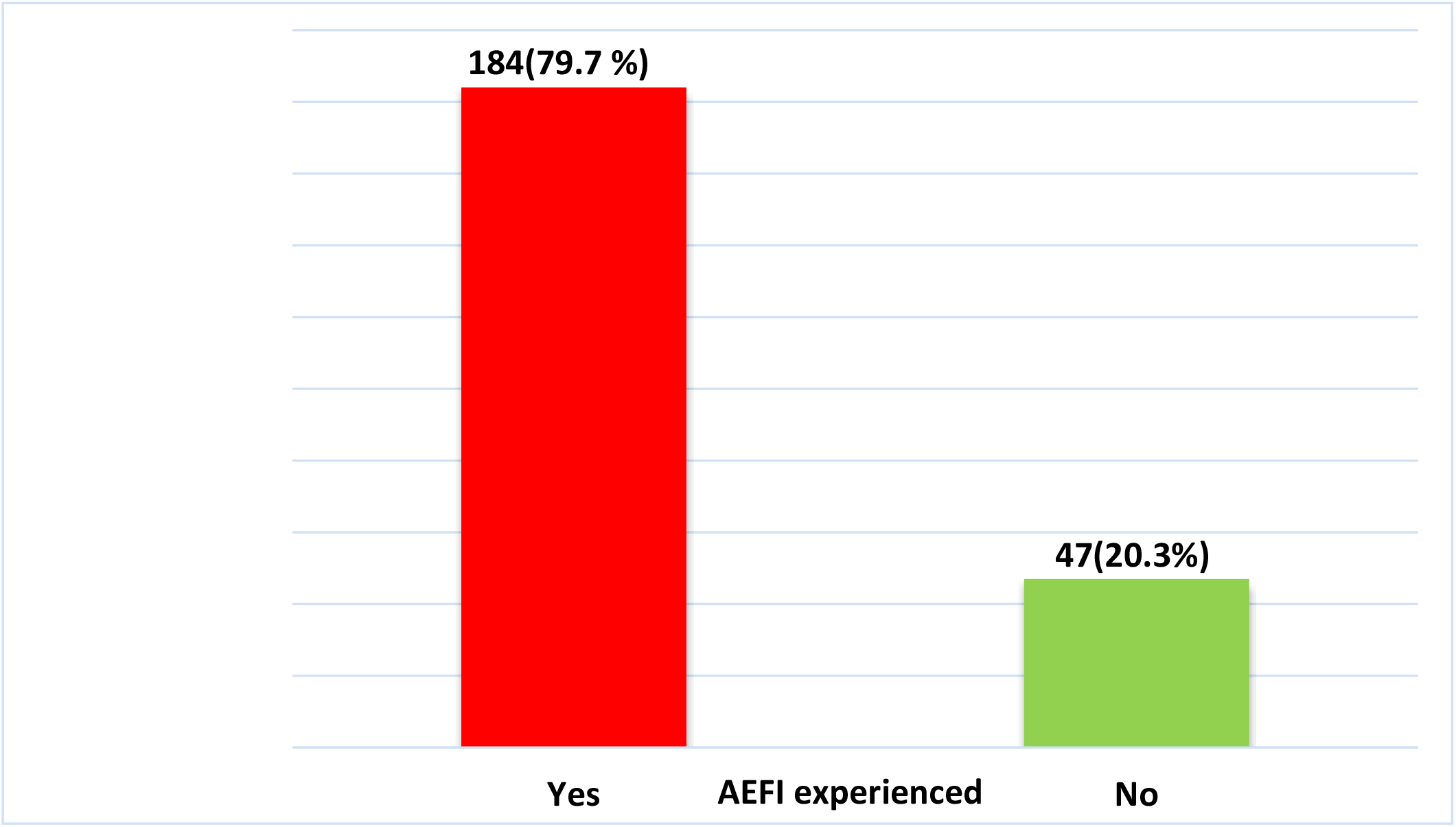
Prevalence of AEFI among respondents. Majority of the participants [184(79.7%)] have experienced AEFI following COVID-19 vaccination (Figure 1).

**FIGURE 2:**
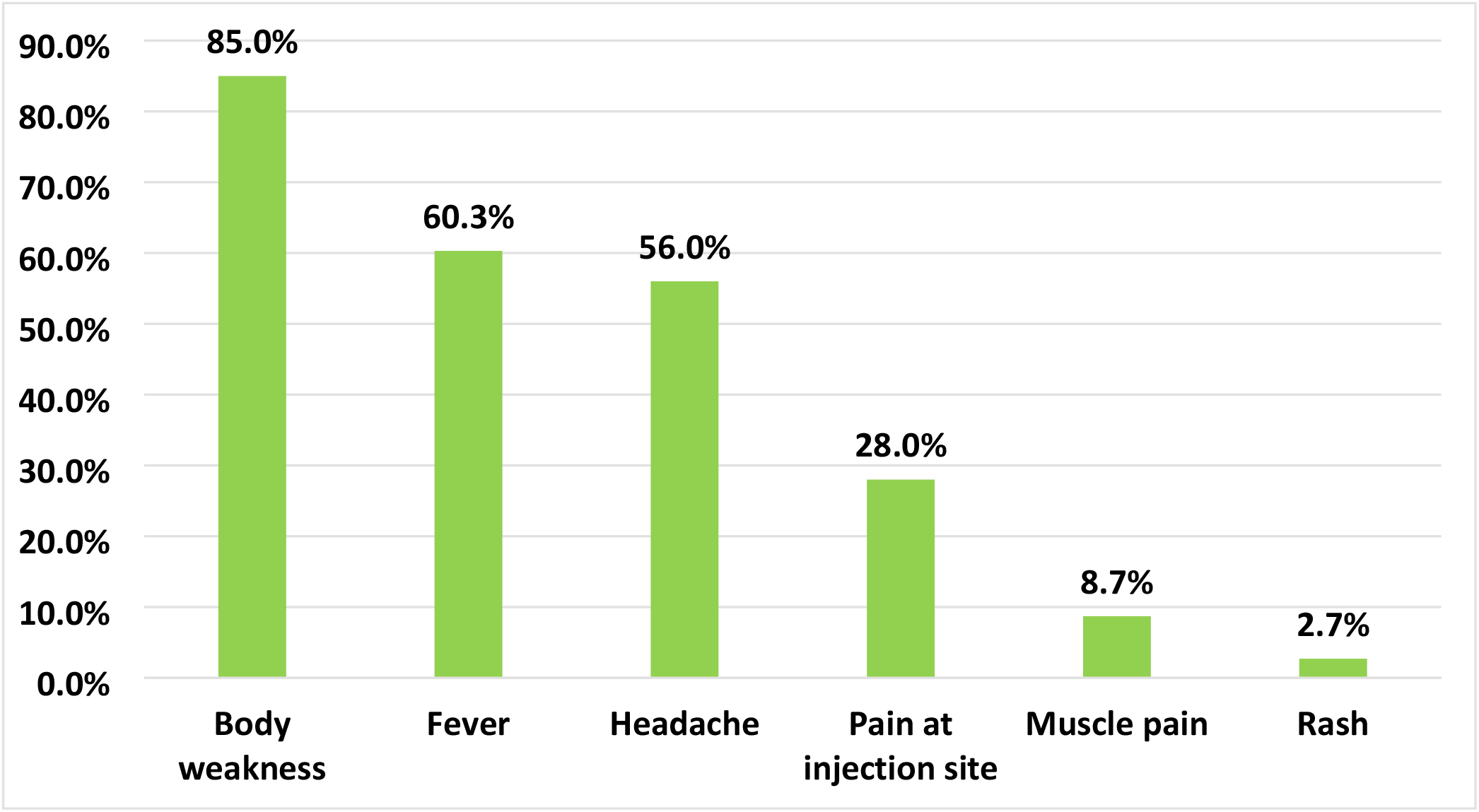
Types of AEFI experienced by the respondents (Multiple options) The most frequent AEFI experienced by the respondents was body weakness (85.0%), followed by fever (60.3%); the least AEFIs experienced by the respondent were rash (2.7%) (Figure 2).

**Table 2:**
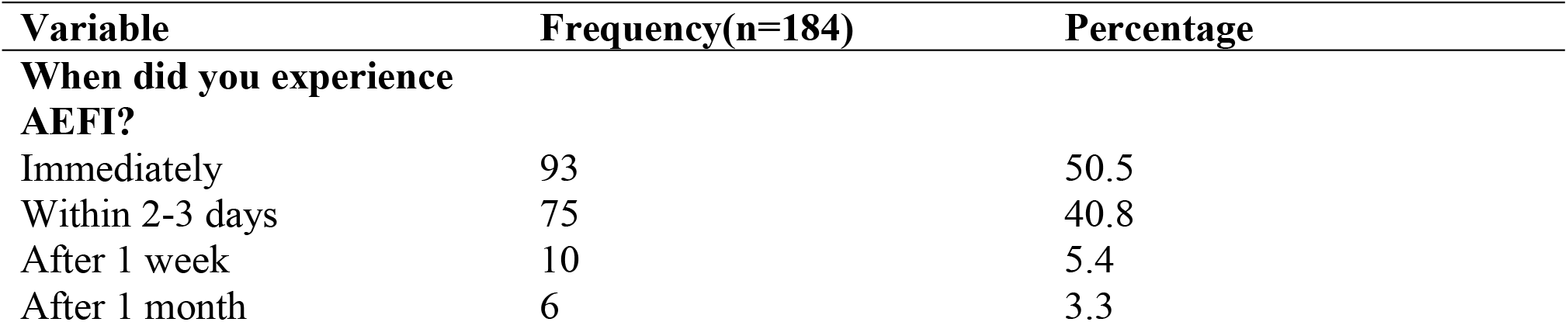
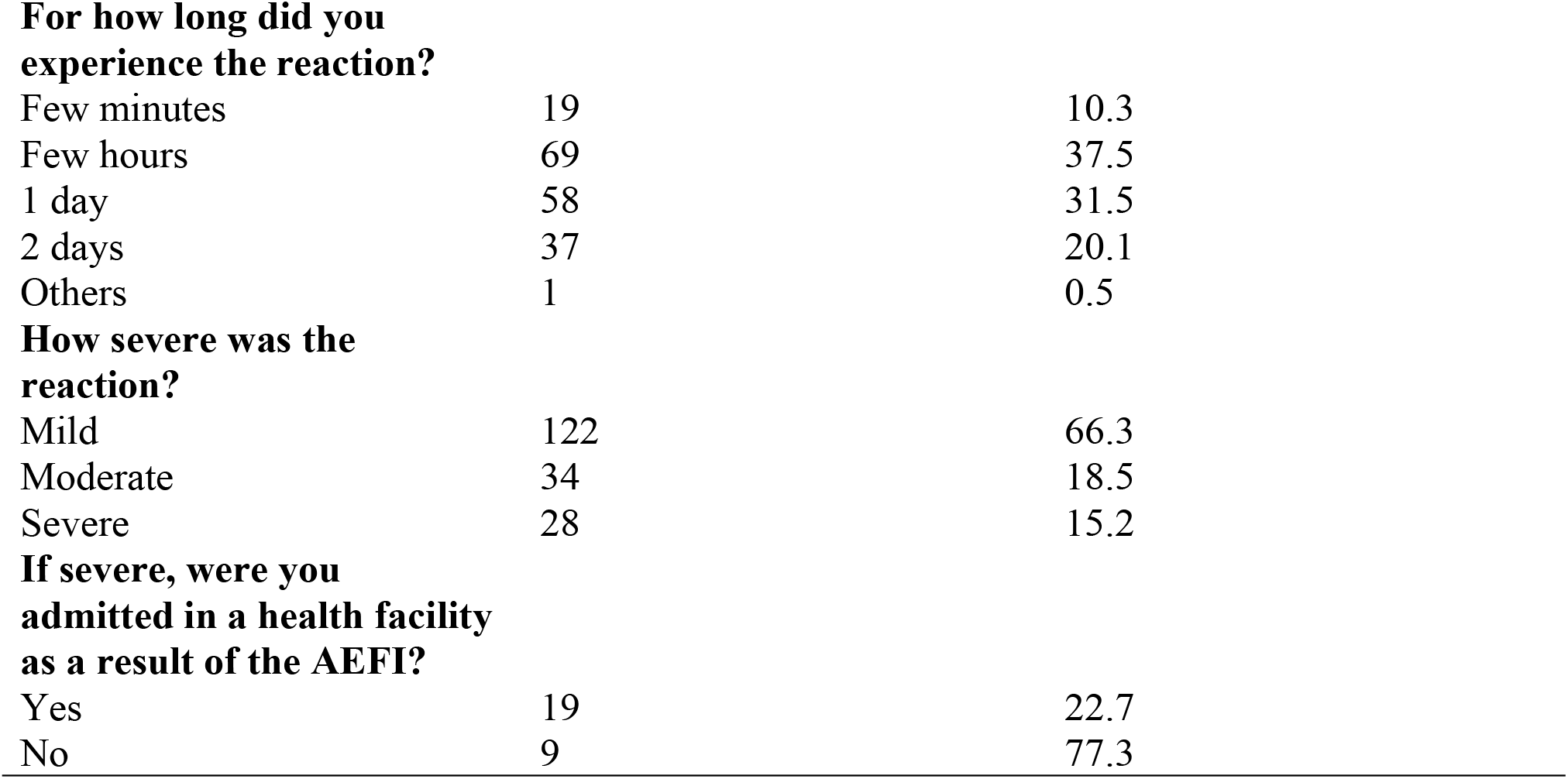
Pattern of AEFIs experienced by the respondents.

| Variable | Frequency(n=184) | Percentage |
| --- | --- | --- |
| **When did you experience AEFI?** |  |  |
| Immediately | 93 | 50.5 |
| Within 2-3 days | 75 | 40.8 |
| After 1 week | 10 | 5.4 |
| After 1 month | 6 | 3.3 |
| **For how long did you experience the reaction?** |  |  |
| Few minutes | 19 | 10.3 |
| Few hours | 69 | 37.5 |
| 1 day | 58 | 31.5 |
| 2 days | 37 | 20.1 |
| Others | 1 | 0.5 |
| **How severe was the reaction?** |  |  |
| Mild | 122 | 66.3 |
| Moderate | 34 | 18.5 |
| Severe | 28 | 15.2 |
| **If severe, were you admitted in a health facility as a result of the AEFI?** |  |  |
| Yes | 19 | 22.7 |
| No | 9 | 77.3 |

Up to half of the respondents that experienced AEFI experienced it immediately (50.5%), whereas 40.8% experienced it within 2-3 days. Most of the reactions lasted between a few hours (37.5%) and one day (35.1%). About two-thirds (66.3%) of the reactions were mild and 15.2% were severe reactions, of which 22.7% required admission to a health facility (Table 2).

**Figure 3:**
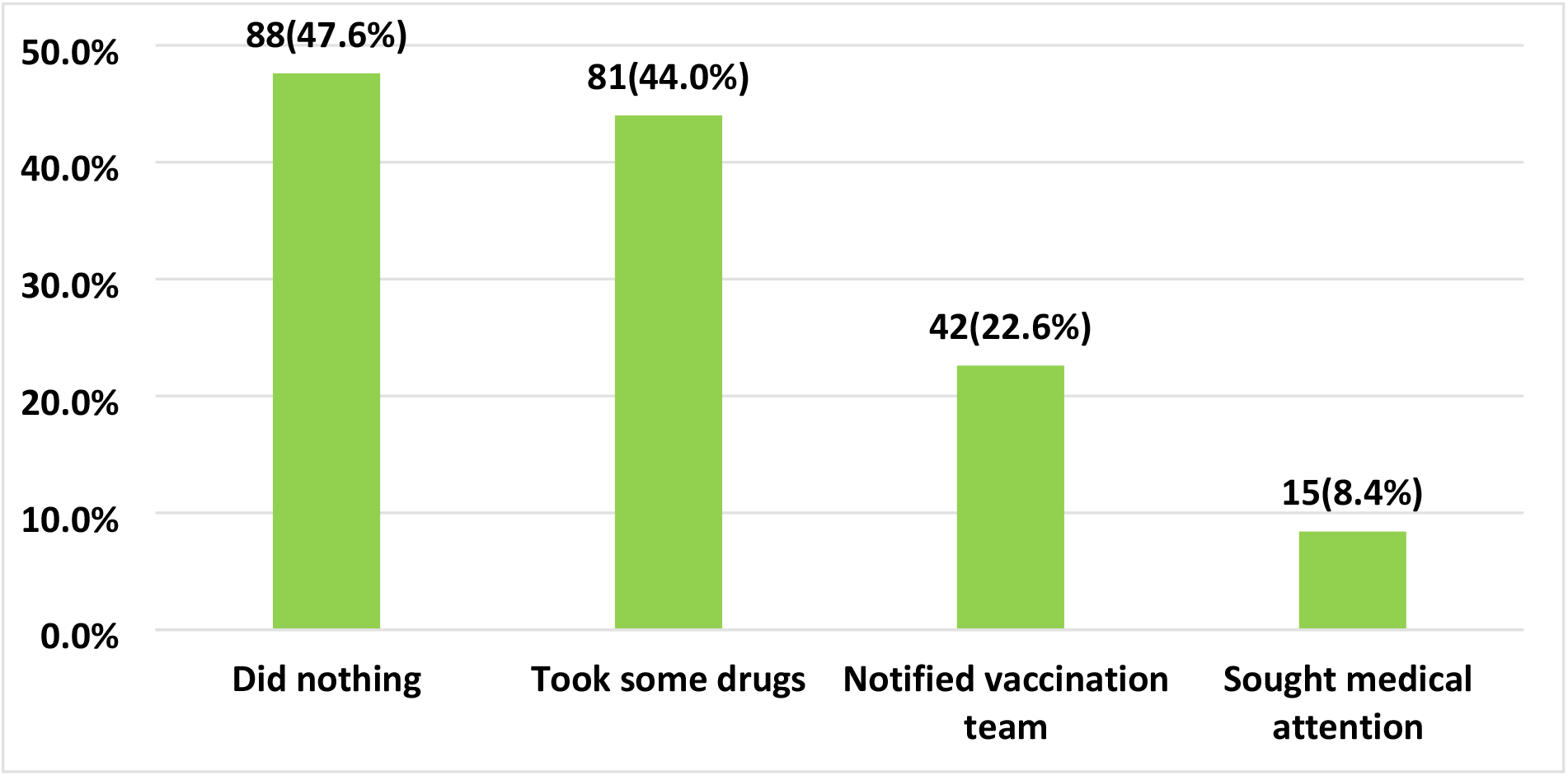
Actions taken after experiencing the AEFI (multiple options) Close to half of the respondents [88(47.6%)] said they did nothing following their experience of AEF; 81(44%) said they took some drugs; and 42(22.6%) said they notified the vaccination about the adverse reaction (Figure 3).

**Table 3.**
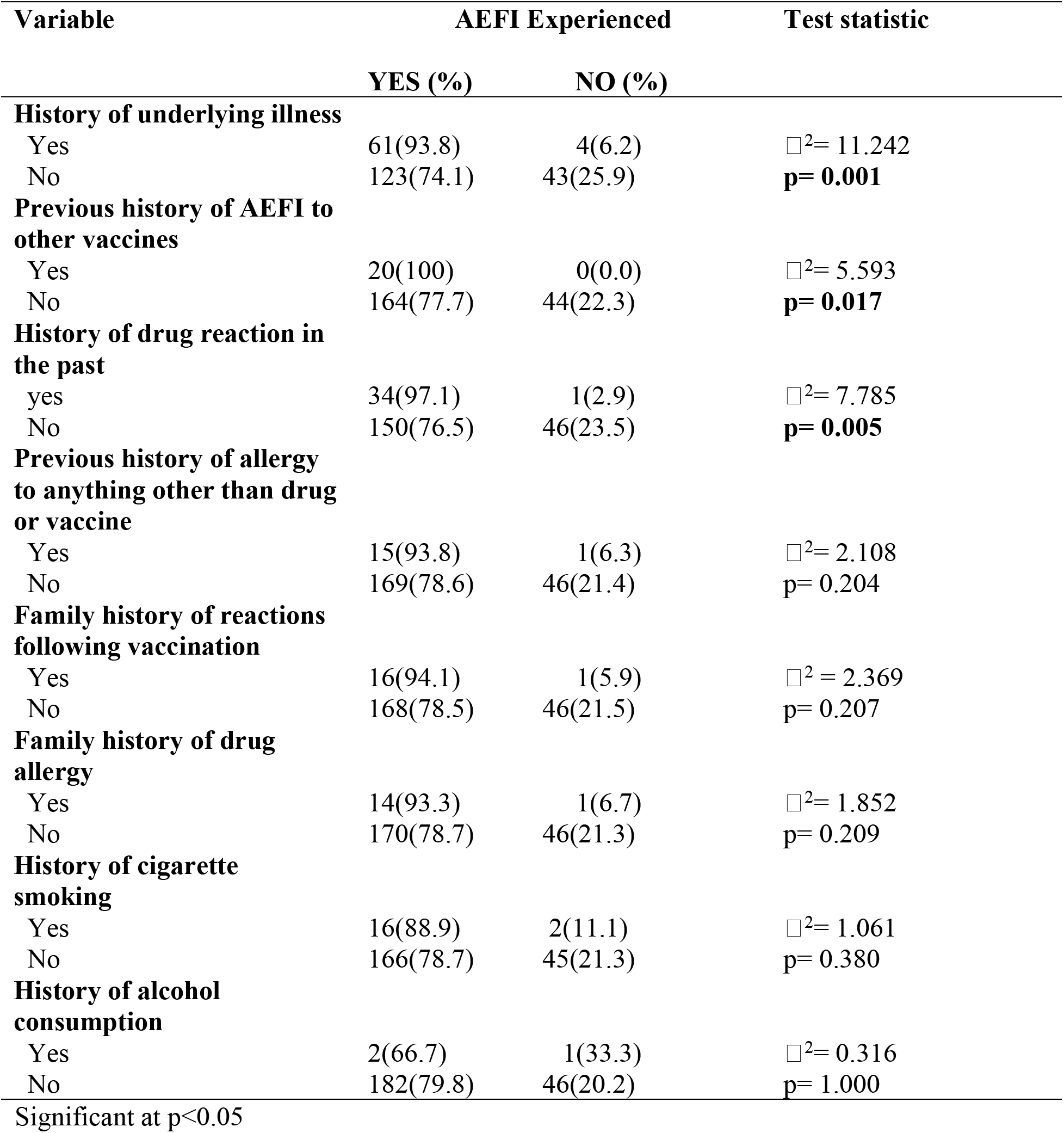
Factors associated with AEFI following COVID-19 vaccination.

AEFIs to COVID-19 vaccine were significantly associated with a history of an underlying medical illness, a previous history of AEFIs to other vaccines, and a previous history of drug reaction (p0.05). Other factors, such as a history of other allergies (other than drugs), cigarette and alcohol use, were not associated with AEFI to COVID-19 vaccines (p>0.05).

The binary logistic regression table (Table 4) shows that those who had a history of an underlying medical condition were about 4 times more likely to have AEFI following COVID-19 vaccination (p = 0.007, OR = 4.448, 95% C I= 1.513-13.077); likewise, those with a previous history of drug reaction were about 10 times more likely to experience the AEFI (p=0.023, OR=10.427, 95% CI=1.389-78.272)

**Table 4:** Binary logistic regression showing the predictors of AEFI following COVID-19 vaccination.

| Predictors | P-value | OR | 95% CI |  |
| --- | --- | --- | --- | --- |
|  |  |  | Lower | Upper |
| History of underlying medical condition |  |  |  |  |
| Yes vs. No* |  |  |  |  |
| History of drug reaction in the past | 0.007 | 4.448 | 1.513 | 13.077 |
| Yes vs. No* | 0.023 | 10.427 | 1.389 | 78.272 |

\*Reference category

## Discussion

This study was conducted to determine the prevalence and pattern of AEFI following COVID-19 vaccination among the adult population in Sokoto metropolis North West, Nigeria.

In this study, up to 79.7% of the respondents have experienced AEFIs following COVID-19 vaccination and this is quite high especially for a vaccine that has recently been introduced. Similar studies conducted in Europe,^18,23^ Asia,^15^ and Africa^16,19^ also reported a high prevalence of adverse reactions to COVID-19. The high prevalence of the adverse reactions observed in these studies is not surprising considering the fact that all vaccines, including the vaccines used in the Expanded Program for Immunization (EPI), have varying forms of adverse reactions.^13,24^ However, the adverse reactions are usually mild (CDC).

Up to two-thirds of the adverse reactions experienced by the respondents in this study were mild, which is also the case for most vaccines.^13,24^ The adverse reactions reported by study participants (body weakness, fever, headache, and pain at injection site) are consistent with previous findings.^17,19,20,25,26^ This pattern and types of adverse reactions have been observed to be more common among recipients of the Astrazeneca vaccine;^14,15.25^ similarly, in this study, all the study participants received only the Astrazeneca vaccine. The side effects of most COVID-19 vaccines usually disappear within a few days of receiving the vaccine;^15,23^ in this study as well, up to 99% of the side effects experienced disappeared within few a minutes to two days. As these side effects are usually mild and quite frequent, it is very important for service providers at the vaccination points to always take time to explain to the recipients of the vaccines that these adverse events may likely occur and that they should not be worried as most of it will resolve within few hours to few days. It is however, important to explain that moderate to severe side effects can also occur^15,19,27^ and in such cases, recipients are usually advised to report to the health facility for further evaluation; in this study, 15.2% of the respondents said they experienced severe adverse events.

It is expected that recipients of the vaccines would report any adverse event experienced after receiving the vaccine; in this study, up to three-quarters of those that experienced an adverse reaction did not report it to the vaccination team. The poor reporting rate observed in this study could have resulted from the fact that the majority of the adverse events experienced by the respondents were mild, so the likelihood of a worsening to severe reaction was perceived to be low. When adverse events are not reported to the appropriate authorities, it makes it more difficult to understand how safe a given vaccine is, unless studies such as this are undertaken. The frequencies of adverse reaction occurring after COVID-19 vaccination differ according to the specific type of vaccine (Astrezenica, BioNtech, Morderna etc) received;^13,18,28^ in this study we could not compare the adverse reactions experienced based on type of vaccine received because as at the time of collecting data for this study, the only vaccine available in all the three health facilities surveyed was the Astrezenica vaccine; therefore, all the study participants received only the Astrazeneca COVID-19 vaccine.

On bivariate analysis of factors associated with AEFI following COVID-19 vaccination, this study revealed that the presence of underlying medical conditions, previous history of reactions to other vaccines and history of drug reactions were statistically significantly associated with development of AEFI following COVID-19 vaccination. On multivariate analysis, however, two factors were found to be significant predictors of AEFI. Those who had a history of an underlying illness were about four times more likely to have AEFI after receiving the COVID-19 vaccine; likewise, those with previous history of drug reactions were about 10 times more likely to experience the AEFI. In a study conducted by Zare et al, it was observed that, prevalence of adverse reactions following COVID-19 vaccination was associated with history of underlying illness.^25^ A study conducted in Vietnam to assess the factors influencing AEFI following COVID-19 vaccination with Oxford Astrazeneca among adult population also reported that underlying medical conditions and previous history of reaction to other vaccines were found to be significantly associated with AEFIs following COVID19 vaccination.^29^

One major limitation of this study is the fact that all the respondents received only the Astrazeneca vaccine, thus it was not possible to compare the rate of adverse events across all the vaccines approved for the COVID-19 vaccination. We also relied on respondents’ self-reported adverse reactions, thus some adverse reactions could have been missed probably because the respondents forgot to mention them. Moreover, some of the respondents were interviewed several weeks after receiving the vaccine. Therefore, their responses could have been affected by some recall bias issues. Nevertheless, we believe their recall was good enough since all the adverse reactions were mentioned to the respondents during the phone call interviews.

## Conclusion

This study revealed that majority of the respondents experienced adverse events following COVID-19 vaccination and the most common AEFIs experienced by respondents were body weakness, fever and headache. The only predictors of adverse reaction following COVID-19 vaccination were the underlying medical condition and a history of drug reaction. As the frequency of adverse reactions is quite high, service providers at each COVID-19 vaccination point should always take time and explain to the recipients of the vaccines that adverse reactions may likely occur, especially among those with an underlying medical condition; they should however, reassure them that most of it usually resolve within few hours to few days. Now that there are up to four different COVID-19 vaccines in use in Sokoto state, there is need for further research to study the different patterns of the adverse reactions for each of the available vaccines

## Data Availability

Data cannot be shared publicly because of ethical confidentiality. Data are available from the researchers Ethics Committee (contact via the Principal researcher at) for researchers who meet the criteria for access to confidential data.

## Notes

### Competing Interest Statement

The authors have declared no competing interest.

### Funding Statement

The author(s) received no specific funding for this work.

### Author Declarations

Research Ehics Committe,Ministry of Health Sokoto state, Nigeria and

